# REACT-1 round 7 updated report: regional heterogeneity in changes in prevalence of SARS-CoV-2 infection during the second national COVID-19 lockdown in England

**DOI:** 10.1101/2020.12.15.20248244

**Authors:** Steven Riley, Caroline E. Walters, Haowei Wang, Oliver Eales, Kylie E. C. Ainslie, Christina Atchison, Claudio Fronterre, Peter J. Diggle, Deborah Ashby, Christl A. Donnelly, Graham Cooke, Wendy Barclay, Helen Ward, Ara Darzi, Paul Elliott

## Abstract

**Background:** England exited a four-week second national lockdown on 2nd December 2020 initiated in response to the COVID-19 pandemic. Prior results showed that prevalence dropped during the first half of lockdown, with greater reductions in higher-prevalence northern regions.

**Methods:** REACT-1 is a series of community surveys of SARS-CoV-2 RT-PCR swab-positivity in England, designed to monitor the spread of the epidemic and thus increase situational awareness. Round 7 of REACT-1 commenced swab-collection on 13th November 2020. A prior interim report included data from 13th to 24th November 2020 for 105,122 participants. Here, we report data for the entire round with swab results obtained up to 3rd December 2020.

**Results:** Between 13th November and 3rd December (round 7) there were 1,299 positive swabs out of 168,181 giving a weighted prevalence of 0.94% (95% CI 0.87%, 1.01%) or 94 per 10,000 people infected in the community in England. This compares with a prevalence of 1.30% (1.21%, 1.39%) from 16th October to 2nd November 2020 (round 6), a decline of 28%. Prevalence during the latter half of round 7 was 0.91% (95% CI, 0.81%, 1.03%) compared with 0.96% (0.87%, 1.05%) in the first half. The national R number in round 7 was estimated at 0.96 (0.88, 1.03) with a decline in prevalence observed during the first half of this period no longer apparent during the second half at the end of lockdown. During round 7 there was a marked fall in prevalence in West Midlands, a levelling off in some regions and a rise in London. R numbers at regional level ranged from 0.60 (0.41, 0.80) in West Midlands up to 1.27 (1.04, 1.54) in London, where prevalence was highest in the east and south-east of the city. Nationally, between 13th November and 3rd December, the highest prevalence was in school-aged children especially at ages 13-17 years at 2.04% (1.69%, 2.46%), or approximately 1 in 50.

**Conclusion:** Between the previous round and round 7 (during lockdown), there was a fall in prevalence of SARS-CoV-2 swab-positivity nationally, but it did not fall uniformly over time or by geography. Continued vigilance is required to reduce rates of infection until effective immunity at the population level can be achieved through the vaccination programme.

## Introduction

In common with other European countries, England has been experiencing an increase in coronavirus cases since September 2020, a so-called second wave [1]. Following a rapid rise in cases through October [2,3], England entered a second national lockdown on 5th November, which continued until 2nd December 2020. The REal-time Assessment of Community Transmission-1 (REACT-1) study has been monitoring the spread of the virus through repeated community-based RT-PCR testing for SARS-CoV-2 based on self-administered throat and nose swabs [4]. These have been obtained from random samples of the population, ranging in size from 100,000 to 170,000, during separate study rounds [2,5–10] that have been carried out approximately monthly since the latter part of the first lockdown in May 2020 [9]. We report here complete results from the seventh round of data collection which took place from 13th November to 3rd December, covering most of the period of the second national lockdown (5th November to 2nd December). Interim results from the seventh round from 13th until 24th November are published elsewhere [2].

## Methods

The methods of the REACT-1 programme are published [4] and study materials and response rates by round are available on our website [11]. Briefly, non-overlapping random samples of the population of England at lower-tier local authority level (LTLA, n=315) are invited to take part in each round of the study based on the National Health Service list of patients. For those registering to take part, a swab kit is sent to a named individual who is requested to provide a self-administered throat and nose swab (or parent/guardian obtains the swab for children aged 12 years or younger). The participant is requested to refrigerate the sample and order a courier for same or next day pick-up and transporting to the laboratory for RT-PCR. Participants complete an online questionnaire (or telephone interview) giving information on history of symptoms, health and lifestyle. Prevalence of SARS-CoV-2 infection is estimated nationally, regionally, sub-regionally, and by socio-demographic and other characteristics. Time trends and reproduction numbers (R) are also estimated, both between subsequent rounds and within rounds using exponential growth models. We provide both unweighted prevalence estimates and estimates weighted to be representative of the population of England as a whole. We use neighbourhood-based spatial smoothing to describe the geographic variation in prevalence by LTLA as previously described [12]. We conduct multivariable logistic regression to estimate odds of swab-positivity according to selected explanatory variables including employment, ethnicity and household size. Statistical analyses are carried out in R [13].

We obtained research ethics approval from the South Central-Berkshire B Research Ethics Committee (IRAS ID: 283787).

## Results

Between 13th November and 3rd December (round 7) there were 1,299 positive swabs out of 168,181 giving a weighted prevalence of 0.94% (95% CI 0.87%, 1.01%) or 94 per 10,000 people. This compares with a prevalence in the previous round (16th October to 2nd November), of 1.30% (1.21%, 1.39%), a fall of 28%. Prevalence during the latter half of round 7 was 0.91% (95% CI, 0.81%, 1.03%) compared with 0.96% (0.87%, 1.05%) in the first half (Table 1).

**Table 1.** Unweighted and weighted prevalence of swab-positivity across seven rounds of REACT-1.

| Round | Tested swabs | Positive swabs | Unweighted prevalence (95% CI) | Weighted prevalence (95% CI) | First sample | Last sample |
| --- | --- | --- | --- | --- | --- | --- |
| 1 | 120,620 | 159 | 0.13% (0.11%, 0.15%) | 0.16% (0.13%, 0.19%) | 1/5/2020 | 1/6/2020 |
| 2 | 159,199 | 123 | 0.077% (0.065%, 0.092%) | 0.088% (0.068%, 0.11%) | 19/6/2020 | 7/7/2020 |
| 3 | 162,821 | 54 | 0.033% (0.025%, 0.043%) | 0.040% (0.027%, 0.053%) | 24/7/2020 | 11/8/2020 |
| 4 | 154,325 | 137 | 0.089% (0.075%, 0.11%) | 0.13% (0.096%, 0.15%) | 20/8/2020 | 8/9/2020 |
| 5 | 174,949 | 824 | 0.47% (0.44%, 0.50%) | 0.60% (0.55%, 0.71%) | 18/9/2020 | 5/10/2020 |
| 6 | 160,175 | 1732 | 1.08% (1.03%, 1.13%) | 1.30% (1.21%, 1.39%) | 16/10/2020 | 2/11/2020 |
| 6a | 85,965 | 863 | 1.00% (0.94%, 1.07%) | 1.28% (1.16%, 1.42%) | 16/10/2020 | 25/10/2020 |
| 6b | 74,210 | 869 | 1.17% (1.10%, 1.25%) | 1.32% (1.20%, 1.45%) | 26/10/2020* | 2/11/2020 |
| 7 | 168,181 | 1299 | 0.77% (0.73%, 0.82%) | 0.94% (0.87%, 1.01%) | 13/11/2020 | 30/11/2020 |
| 7a | 105,122 | 821 | 0.78% (0.73%, 0.84%) | 0.96% (0.87%, 1.05%) | 13/11/2020 | 24/11/2020 |
| 7b | 63,059 | 478 | 0.76% (0.69%, 0.83%) | 0.91% (0.81%, 1.03%) | 25/11/2020* | 3/12/2020 |

\*Includes some samples from prior period

From the start of round 6 on 16th October to the end of round 7 on 3rd of December, we estimated an average national R number less than 1 at 0.94 (0.92, 0.95) (Table 2, Figure 1). However, the substantial decline in prevalence observed during the first half of round 7 [2] was no longer apparent in the second half (Figure 2). Overall within round 7, we estimated the national R number to be 0.96 (0.88, 1.03) (Table 2).

**Table 2.** Estimates of national growth rates, doubling times and reproduction numbers for round 7 (7a and 7b combined), rounds 6b and 7a, and rounds 6 (6a and 6b combined) and 7 (7a and 7b combined).

| Round | Outcome | Growth rate | R | Probability R>1 | Halving(-) / Doubling(+) time |
| --- | --- | --- | --- | --- | --- |
| 7 (a and b combined) | All positives | -0.007 ( -0.020 , 0.005 ) | 0.96 ( 0.88 , 1.03 ) | 0.12 | * ( -35.4 , * ) |
|  | Non-symptomatics | -0.004 ( -0.025 , 0.016 ) | 0.97 ( 0.85 , 1.10 ) | 0.34 | * ( -28.1 , 44.3 ) |
|  | Positive for both E and N genes | -0.007 ( -0.021 , 0.008 ) | 0.96 ( 0.87 , 1.05 ) | 0.19 | * ( -32.7 , * ) |
|  | Positive for both E and N genes or positive only for N gene with CT 35 or less | -0.006 ( -0.019 , 0.007 ) | 0.96 ( 0.88 , 1.05 ) | 0.18 | * ( -36.3 , * ) |
| 6b & 7a | All positives | -0.019 ( -0.023 , -0.015 ) | 0.88 ( 0.86 , 0.91 ) | <0.01 | -36.7 ( -30.2 , -46.9 ) |
|  | Non-symptomatics | -0.028 ( -0.034 , -0.021 ) | 0.83 ( 0.80 , 0.87 ) | <0.01 | -25.2 ( -20.3 , -33.1 ) |
|  | Positive for both E and N genes | -0.017 ( -0.022 , -0.012 ) | 0.90 ( 0.87 , 0.93 ) | <0.01 | -41.0 ( -31.8 , * ) |
|  | Positive for both E and N genes or positive only for N gene with CT 35 or less | -0.017 ( -0.021 , -0.012 ) | 0.90 ( 0.87 , 0.92 ) | <0.01 | -41.4 ( -32.7 , * ) |
| 6 (a and b combined) & 7 (a and b combined) | All positives | -0.010 ( -0.012 , -0.008 ) | 0.94 ( 0.92 , 0.95 ) | <0.01 | * ( * , * ) |
|  | Non-symptomatics | -0.018 ( -0.021 , -0.014 ) | 0.89 ( 0.87 , 0.91 ) | <0.01 | -39.1 ( -32.3 , -49.5 ) |
|  | Positive for both E and N genes | -0.008 ( -0.010 , -0.005 ) | 0.95 ( 0.94 , 0.97 ) | <0.01 | * ( * , * ) |
|  | Positive for both E and N genes or positive only for N gene with CT 35 or less | -0.008 ( -0.011 , -0.006 ) | 0.95 ( 0.93 , 0.96 ) | <0.01 | * ( * , * ) |

\* Doubling/Halving time had an estimated magnitude greater than 50 days and so represented approximately constant prevalence

**Figure 1.**
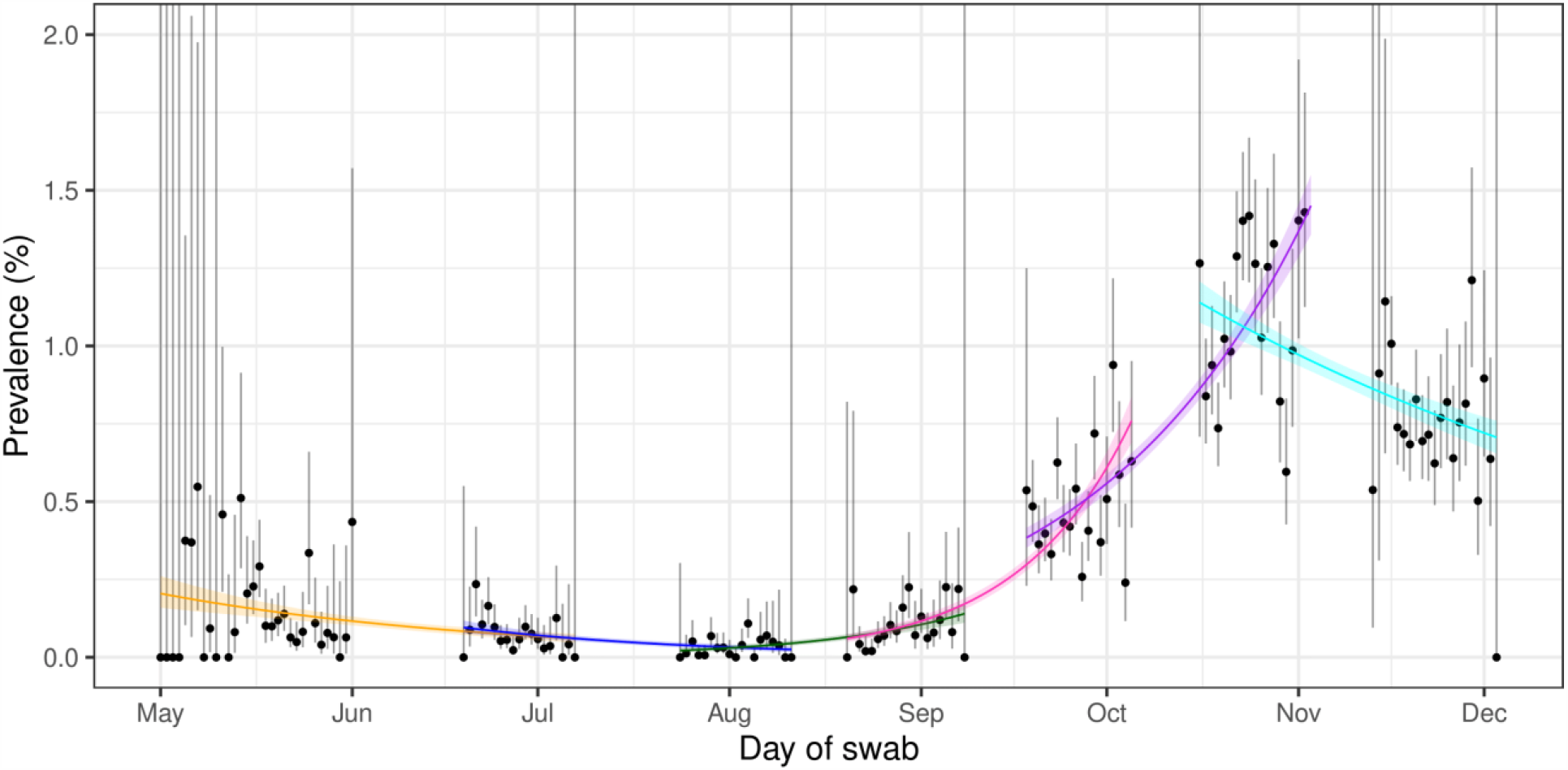
Constant growth rate models fit to REACT-1 data for England for sequential rounds; 1 and 2 (yellow), 2 and 3 (blue), 3 and 4 (green), 4 and 5 (pink), 5 and 6 (purple), and 6 and 7 (cyan).

**Figure 2.**
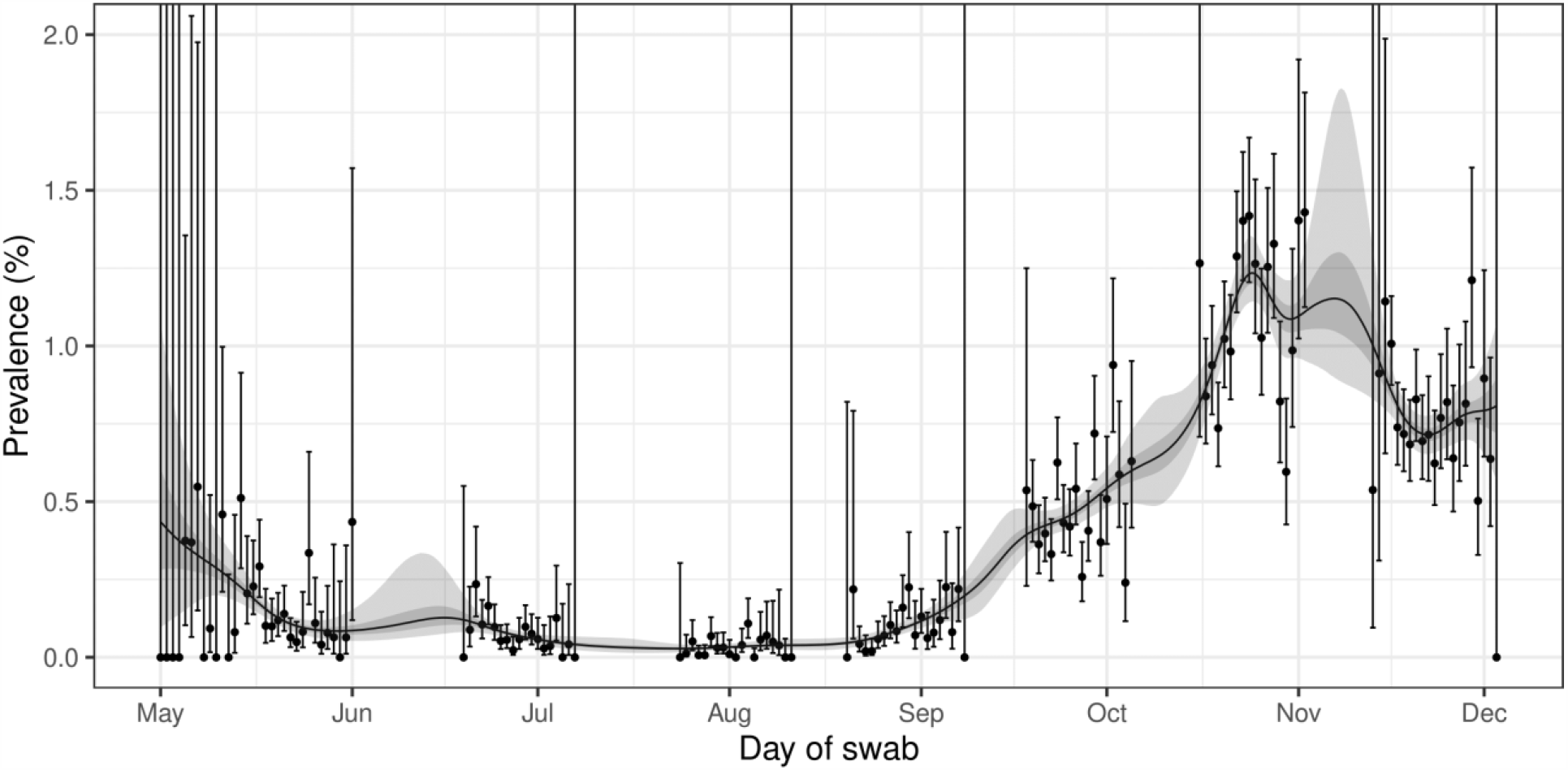
Prevalence of national swab-positivity for England estimated using a p-spline for the full period of the study with central 50% (dark grey) and 95% (light grey) posterior credible intervals.

Between the first and second halves of round 7, there were apparent changes in swab-positivity rates at the regional scale, but with substantial overlaps in confidence intervals (Table 3a, Figure 3). Prevalence in Yorkshire and The Humber was the highest in the second half of round 7, having risen from 1.17% (0.87%, 1.56%) to 1.39% (0.93%, 2.07%). There was also a rise in prevalence in London from 0.98% (0.75%, 1.28%) to 1.21% (0.91%, 1.59%) and in the North East from 0.72% (0.42%, 1.24%) to 1.26% (0.78%, 2.04%). There was a halving of prevalence in West Midlands from 1.55% (1.14%, 2.10%) to 0.71% (0.43%, 1.16%) and a fall in East Midlands from 1.27% (1.03%, 1.57%) to 1.04% (0.79%, 1.38%), and in North West from 1.08% (0.86%, 1.35%) to 0.92% (0.63%, 1.33%).

**Table 3a.**
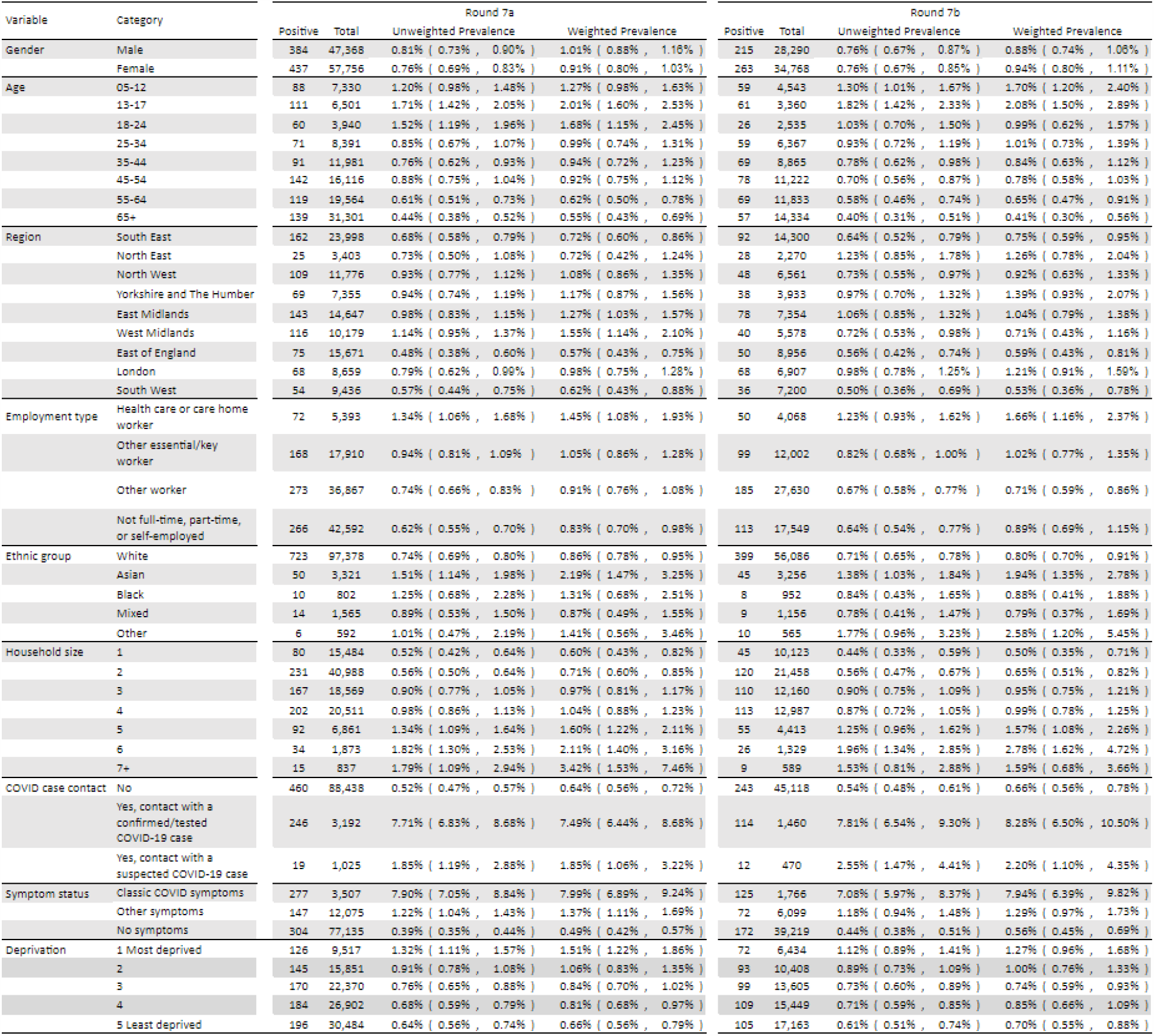
Unweighted and weighted prevalence of swab-positivity for rounds 7a and 7b.

**Table 3b.**
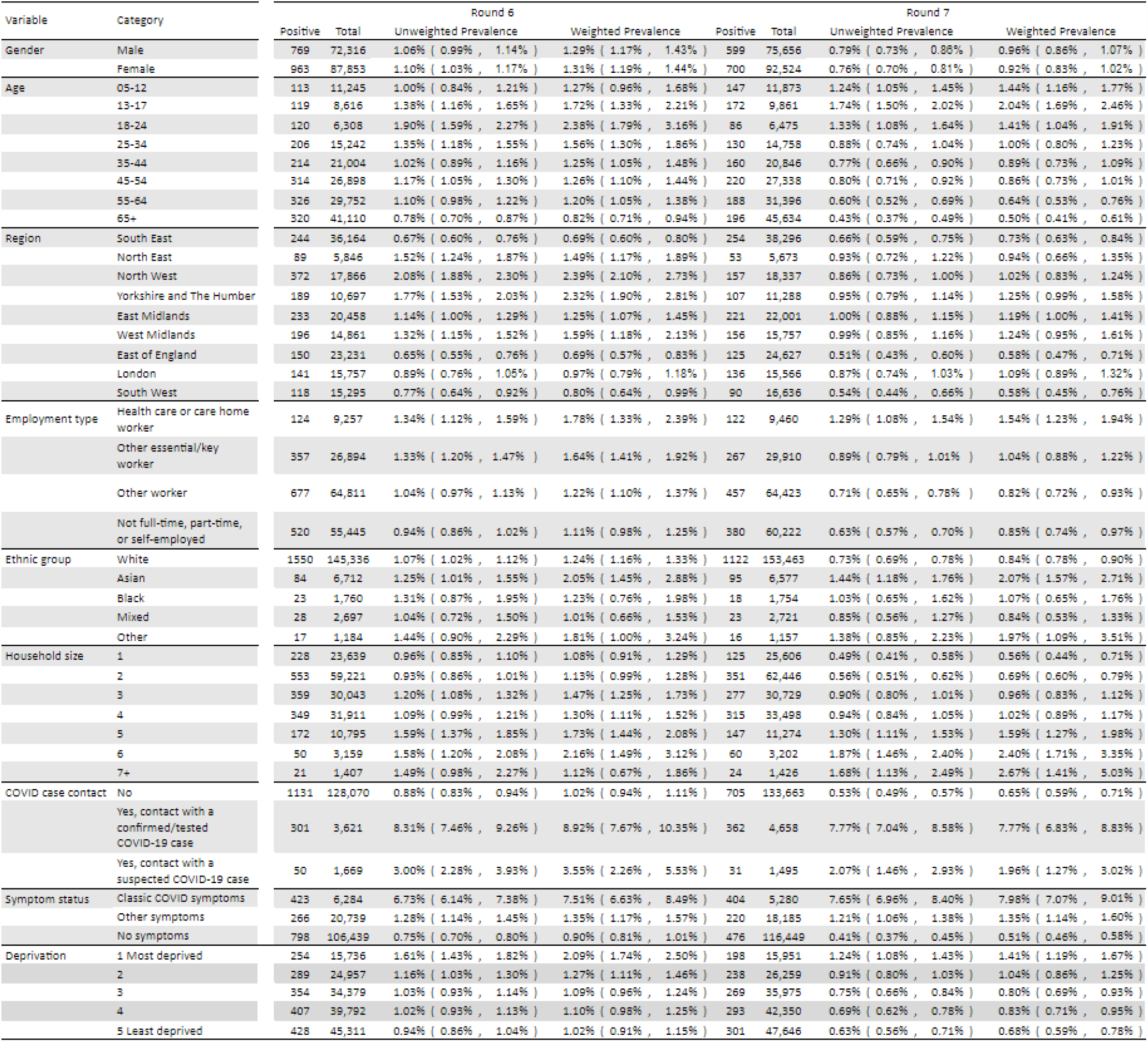
Unweighted and weighted prevalence of swab-positivity for rounds 6 and 7.

**Figure 3.**
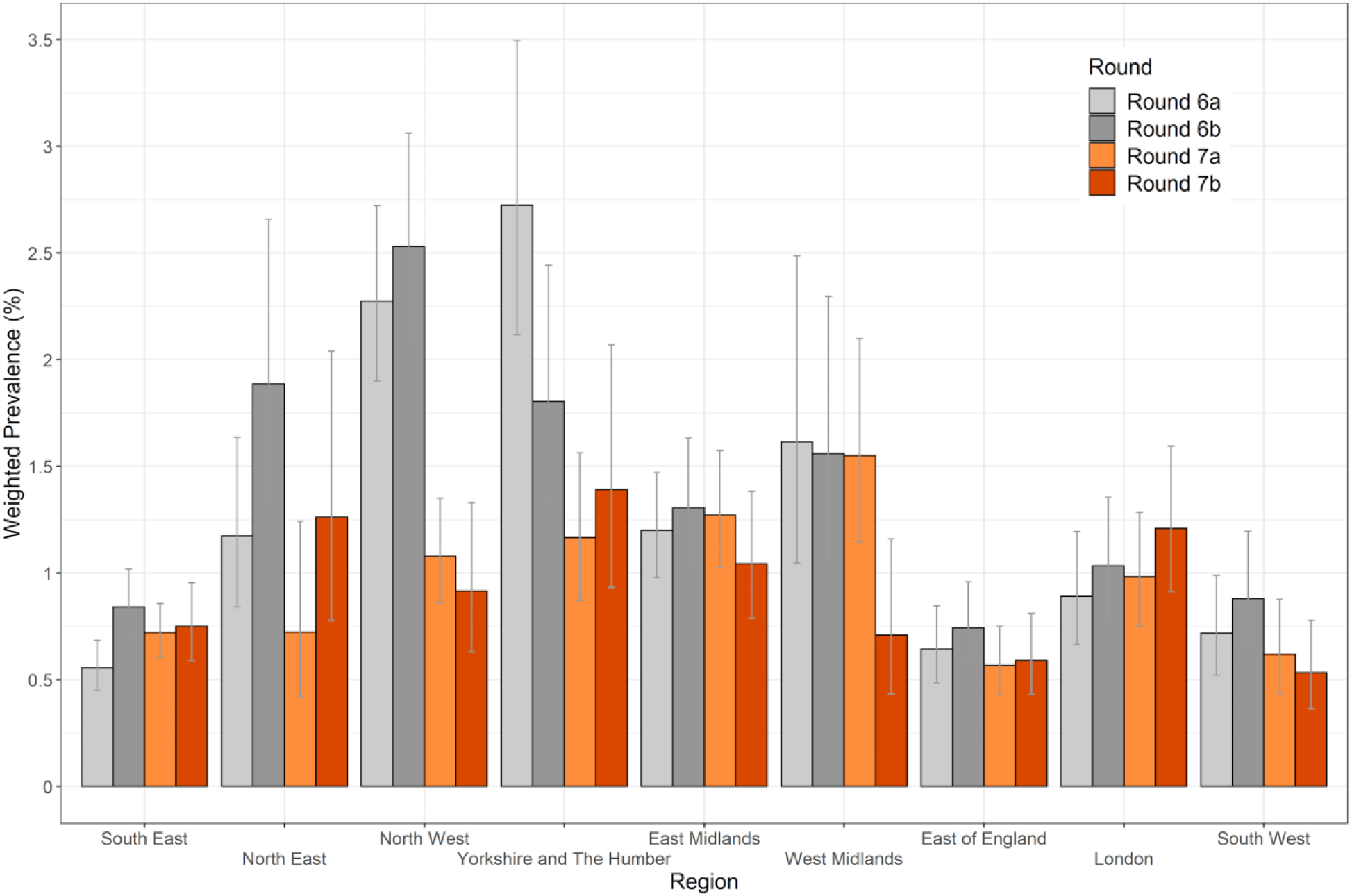
Weighted prevalence of swab-positivity by region for rounds 6a, 6b, 7a, and 7b. Bars show 95% confidence intervals.

At the regional level, R numbers for round 7 overall ranged from 0.60 (0.41, 0.80) for the West Midlands up to 1.27 (1.04, 1.54) for London (Table 4). Also, splines fit to regional prevalence using daily data from the entire REACT-1 dataset can detect short-term recent trends, albeit with high uncertainty (Figure 4). They show a marked reduction in rates in North West and West Midlands but evidence for a flattening off or recent rise in South East and East of England and a rise in London.

**Table 4.**
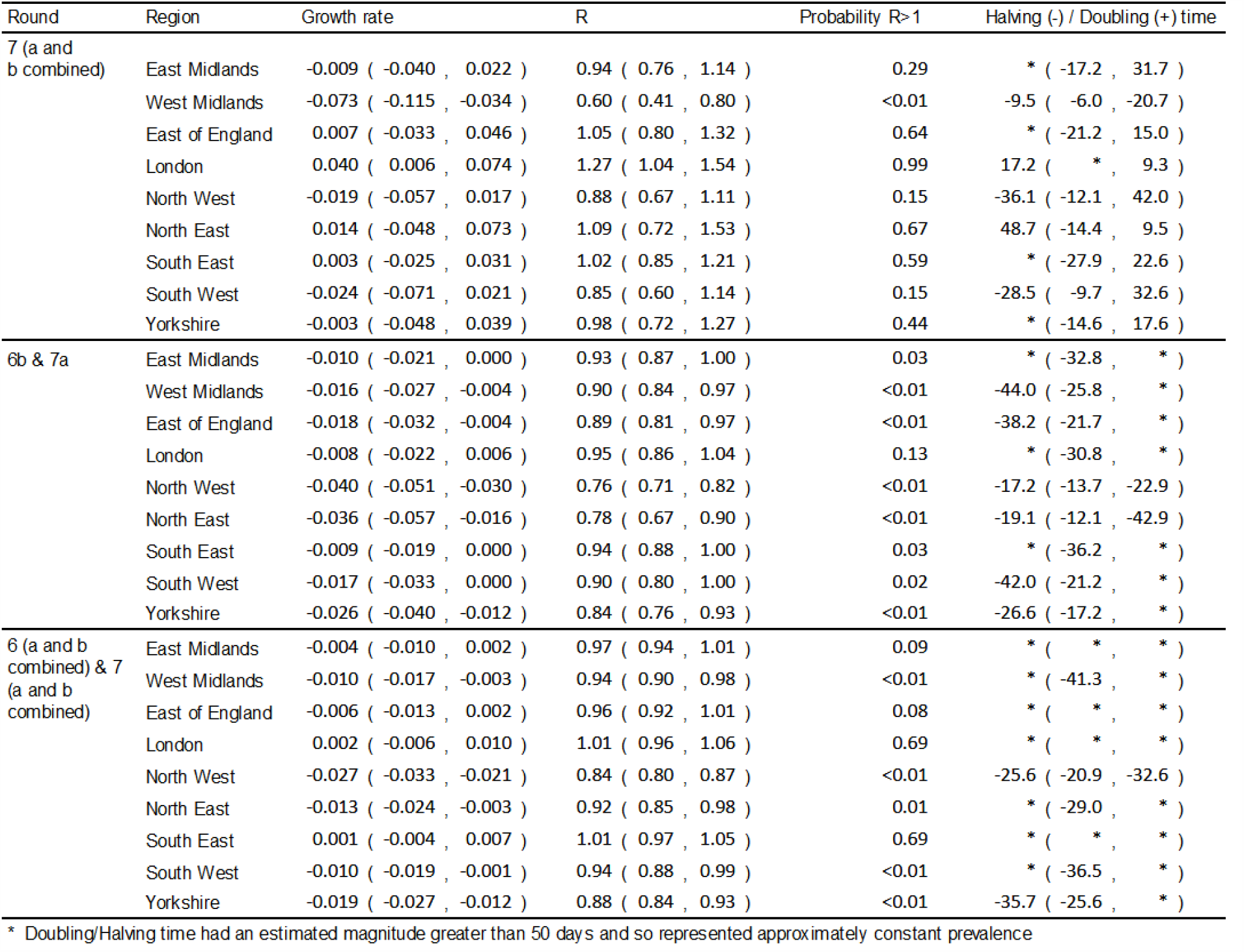
Estimates of regional growth rates, doubling times and reproduction numbers for round 7 (7a and 7b combined), rounds 6b and 7a, and rounds 6 (6a and 6b combined) and 7 (7a and 7b combined).

**Figure 4.**
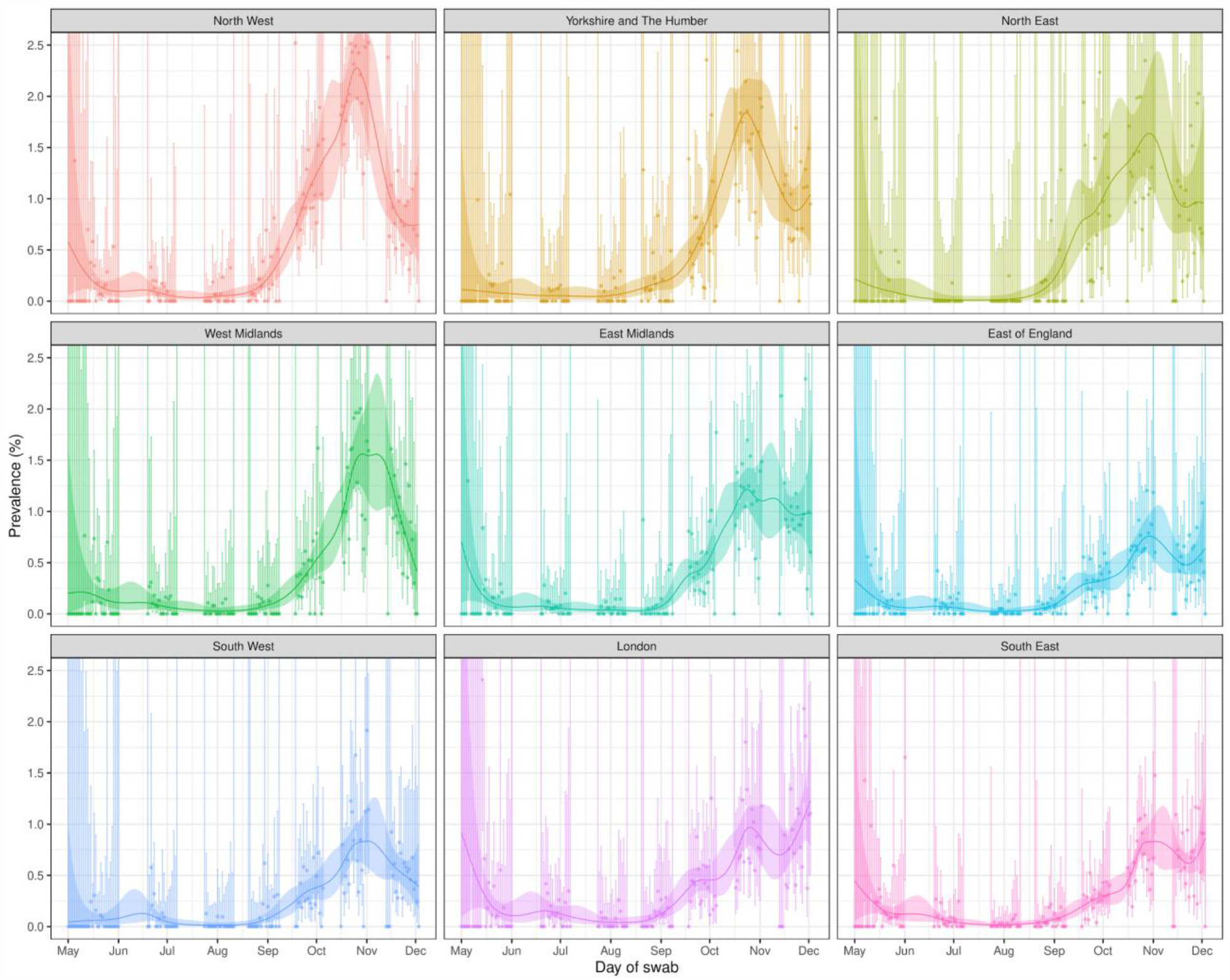
Prevalence of swab-positivity for each region estimated using a p-spline (with a constant second-order random walk prior) for the full period of the study with central 95% posterior credible intervals.

At the sub-regional level there was countrywide heterogeneity in prevalence (Figure 5) and also within London, with highest prevalence in the east and south-east of the city, where prevalence appears to be increasing most (Figure 6). Comparing the first and second halves of round 7 in other regions of England, there were local areas in each region where prevalence rates appeared to be increasing, especially in the east of the country (Figure 7) including areas in East of England (Essex) and South East (Kent) adjacent to London.

**Figure 5.**
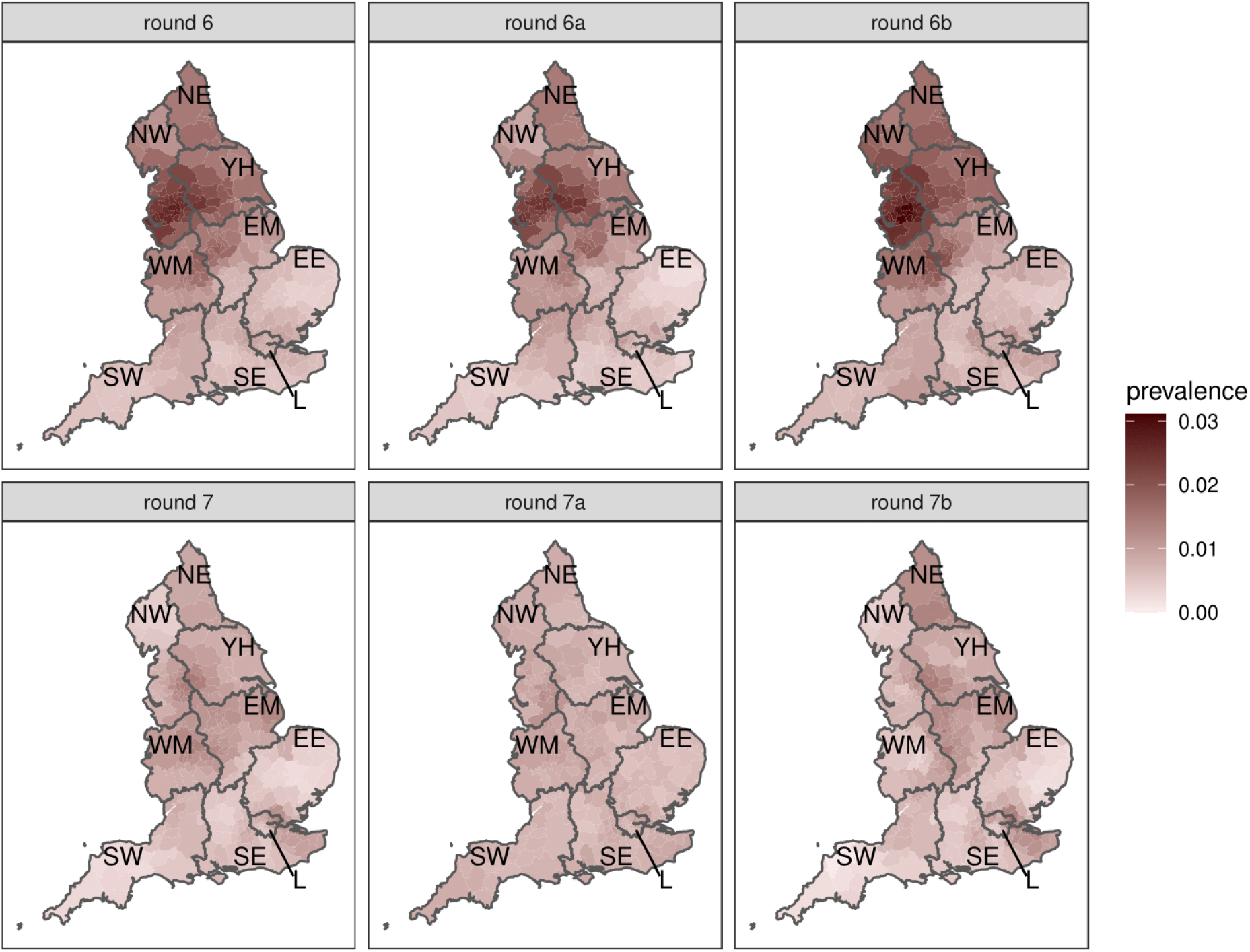
Neighbourhood prevalence for rounds 6 (6a and 6b combined), 6a, 6b, 7 (7a and 7b combined), 7a, and 7b. Neighbourhood prevalence calculated from nearest neighbours (the median number of neighbours within 30 km in the study). Average neighbourhood prevalence displayed for individual lower tier local authorities. Regions: NE = North East, NW = North West, YH = Yorkshire and The Humber, EM = East Midlands, WM = West Midlands, EE = East of England, L = London, SE = South East, SW = South West. Data for unweighted point estimate of prevalence available in the supplementary data file.

**Figure 6.**
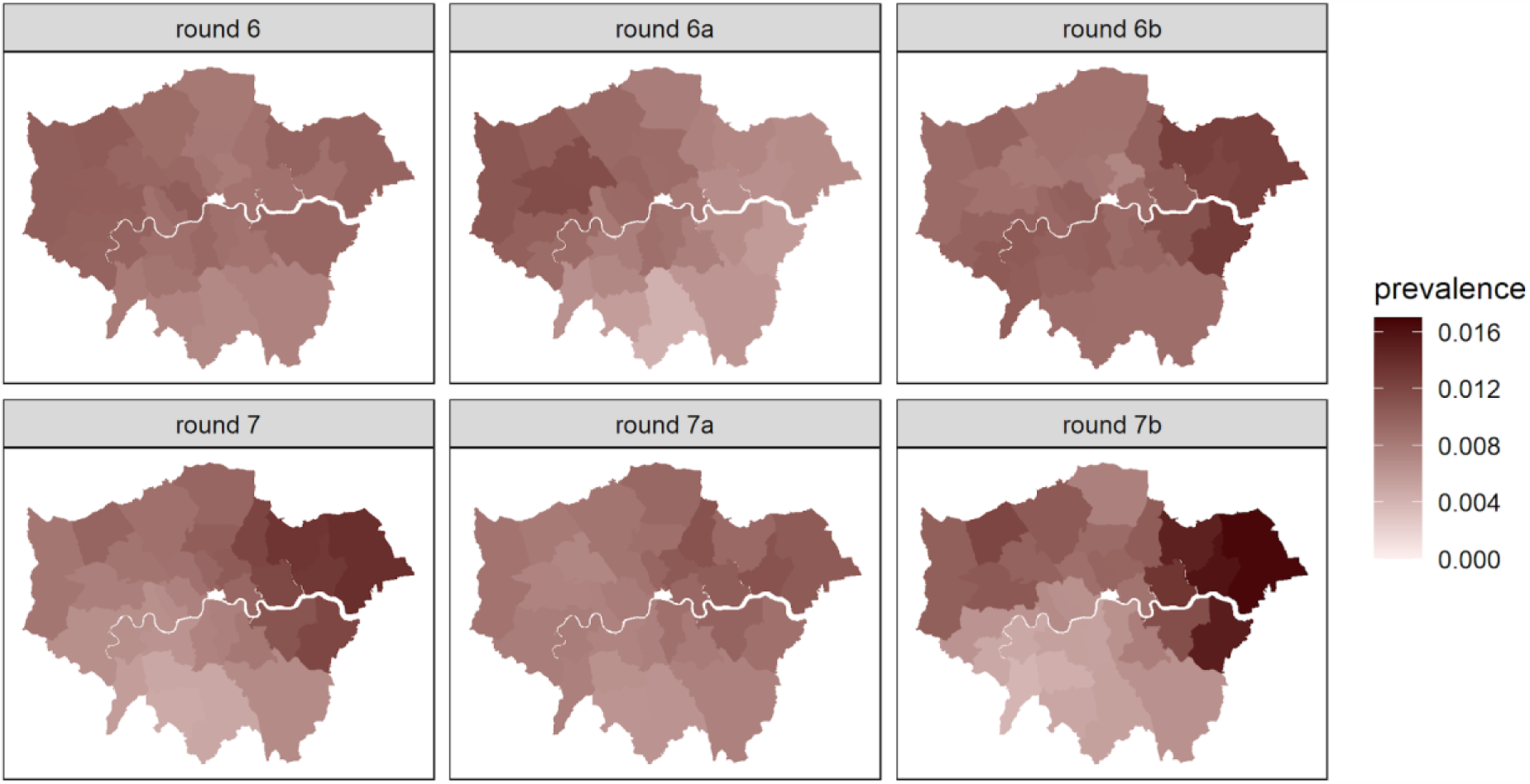
Neighbourhood prevalence for rounds 6 (6a and 6b combined), 6a, 6b, 7 (7a and 7b combined), 7a, and 7b for lower-tier local authorities in London. Neighbourhood prevalence calculated from nearest neighbours (the median number of neighbours within 30 km in the study). Average neighbourhood prevalence displayed for individual lower tier local authorities.

**Figure 7.**
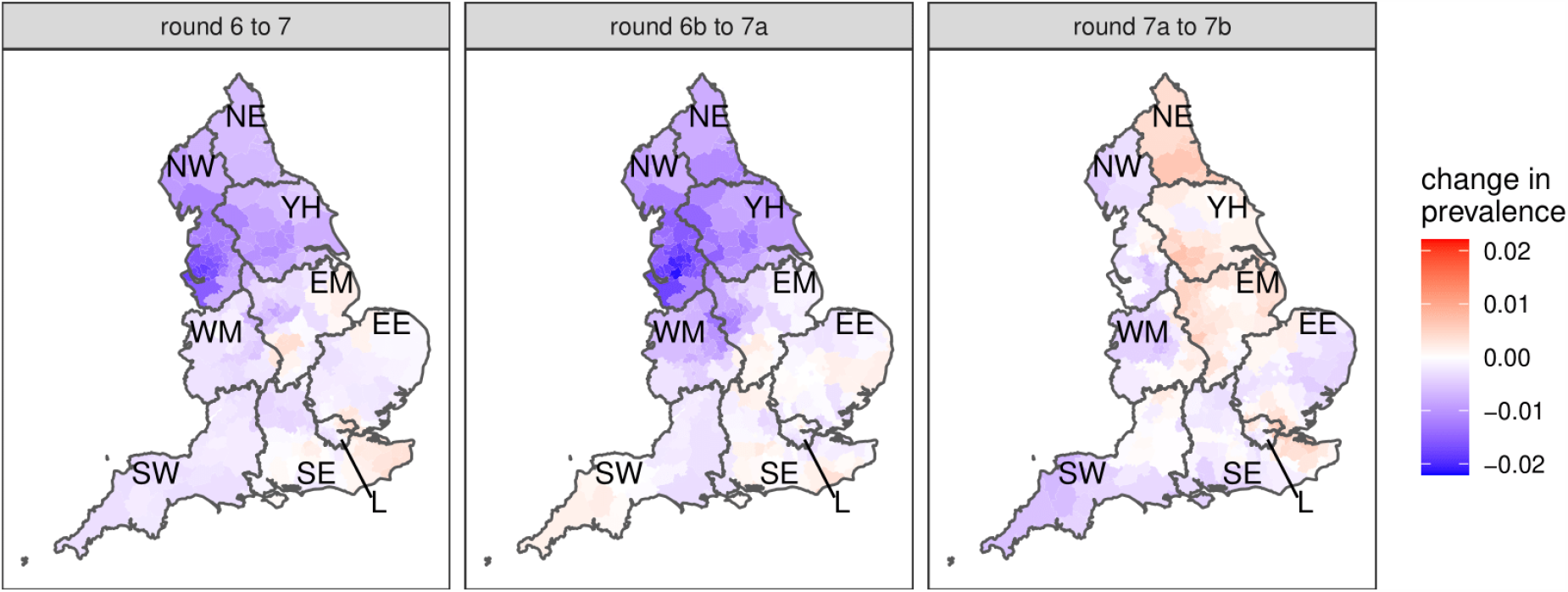
Difference in neighbourhood prevalence at lower tier local authority level from: round 6 to 7; 6b to 7a; and 7a to 7b. Neighbourhood prevalence calculated from nearest neighbours (the median number of neighbours within 30 km in the study). Average neighbourhood prevalence and difference displayed for individual lower tier local authorities. Regions: NE = North East, NW = North West, YH = Yorkshire and The Humber, EM = East Midlands, WM = West Midlands, EE = East of England, L = London, SE = South East, SW = South West.

Patterns of national weighted prevalence by age group show highest levels in school-aged children (Table 3a, Table 3b, Figure 8). At ages 13 to 17 years prevalence in the second half of round 7 was 2.08% (1.50%, 2.89%) and at ages 5 to 12 years it is 1.70% (1.20%, 2.40%) (Table 3a). Nationally, there were large falls in prevalence in 18 to 24 year olds (Table 3a, Table 3b). Splines fit to these age patterns support the trends in younger people and also show decline and then levelling off at ages 55 years and above (Figure 9).

**Figure 8.**
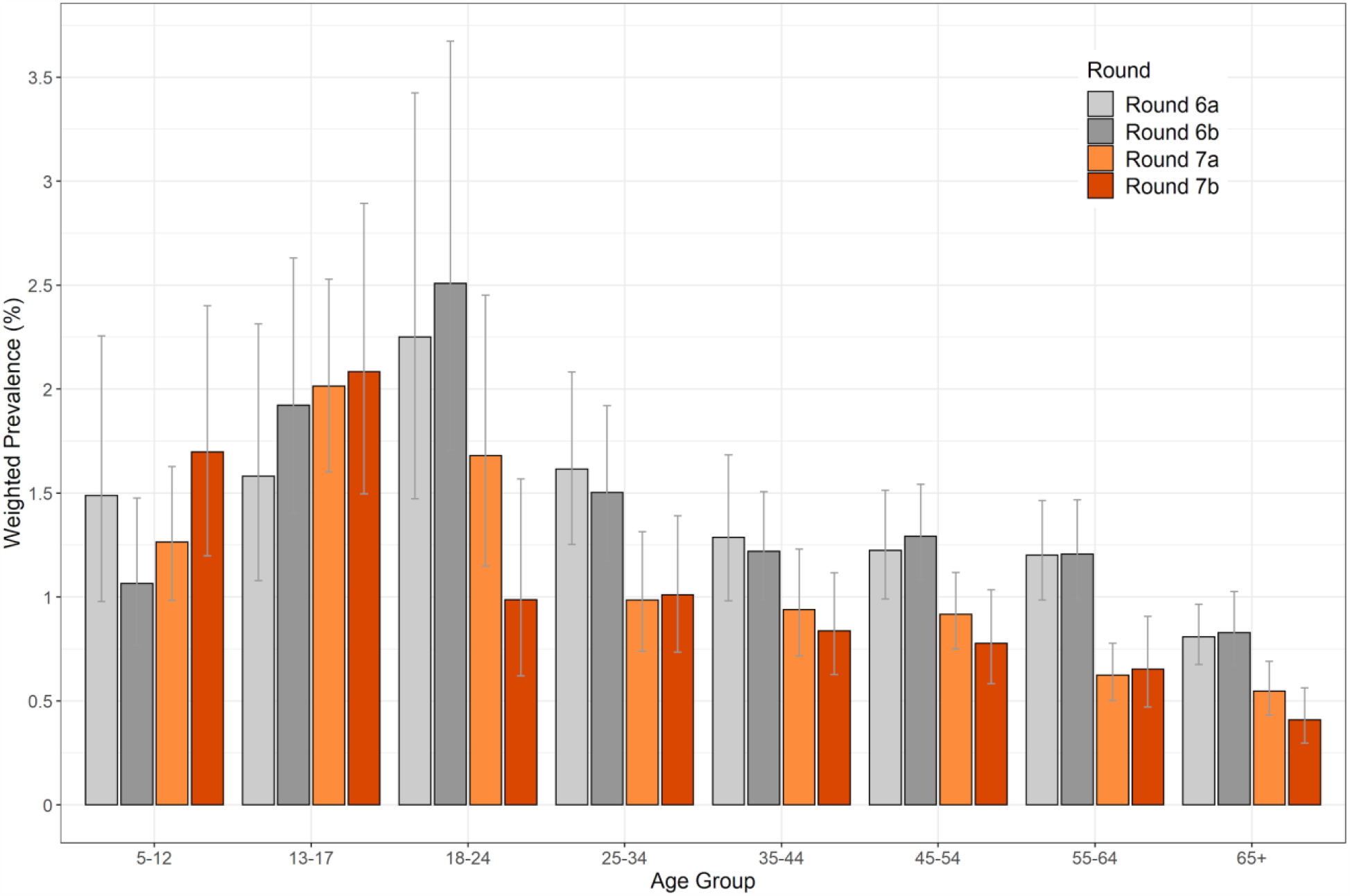
Weighted prevalence of swab-positivity by age group for rounds 6a, 6b, 7a, and 7b. Bars show 95% confidence intervals.

**Figure 9.**
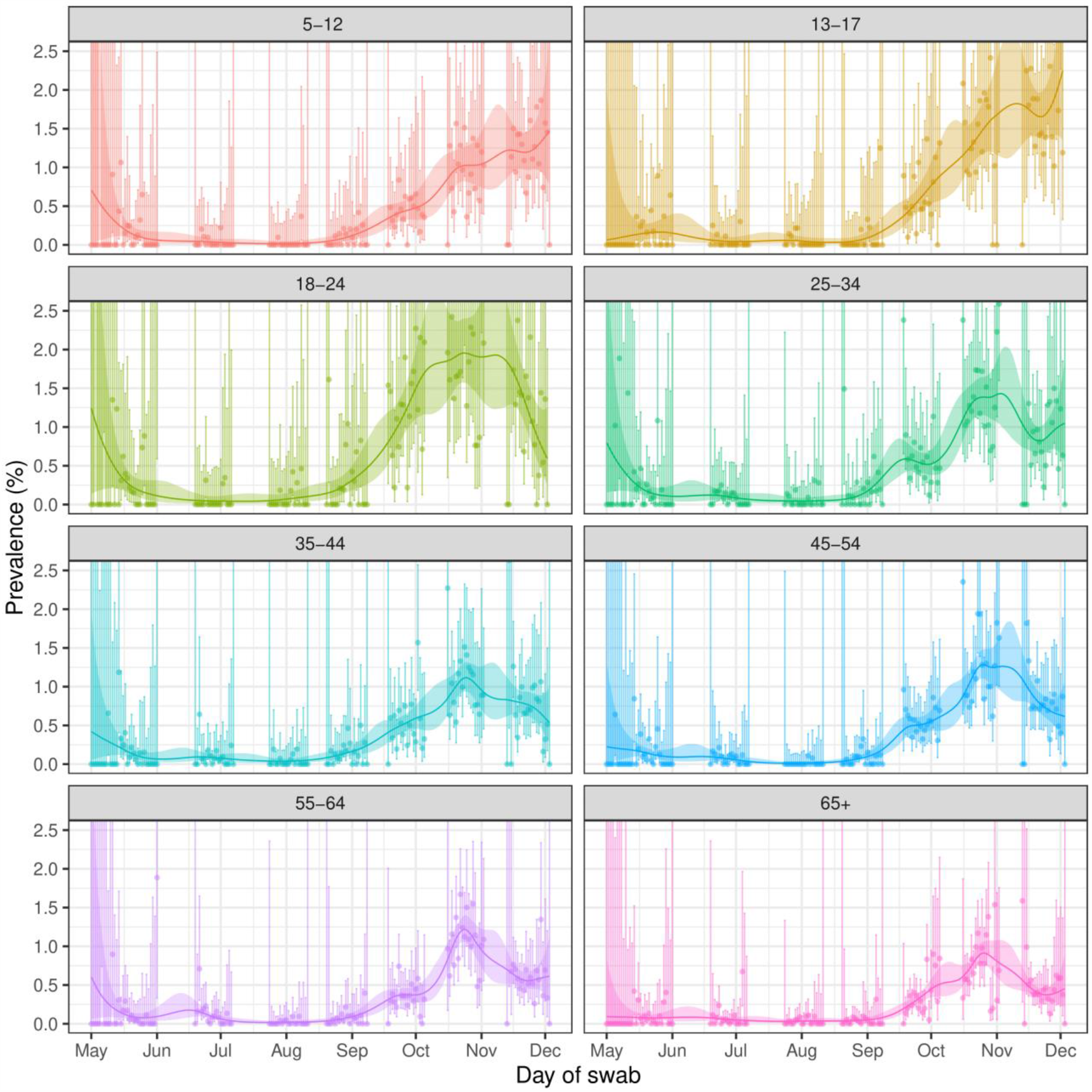
Prevalence of swab-positivity for each age group estimated using a p-spline (with a constant second-order random walk prior) for the full period of the study with central 95% posterior credible intervals.

Patterns of swab-positivity by ethnicity and key worker status were similar in the first and second halves of round 7 (Figure 10). In the second half of round 7, prevalence of swab-positivity in people of Asian and Other ethnicities was 1.94% (1.35%, 2.78%) and 2.58% (1.20%, 5.45%) respectively, compared with 0.80% (0.70%, 0.91%) in white people, while prevalence in health care workers and care home workers was 1.66% (1.16%, 2.37%) compared with 0.71% (0.59%, 0.86%) in other workers (Table 3a). Odds ratios from a multivariable logistic regression model support these findings (Figure 10).

**Figure 10.**
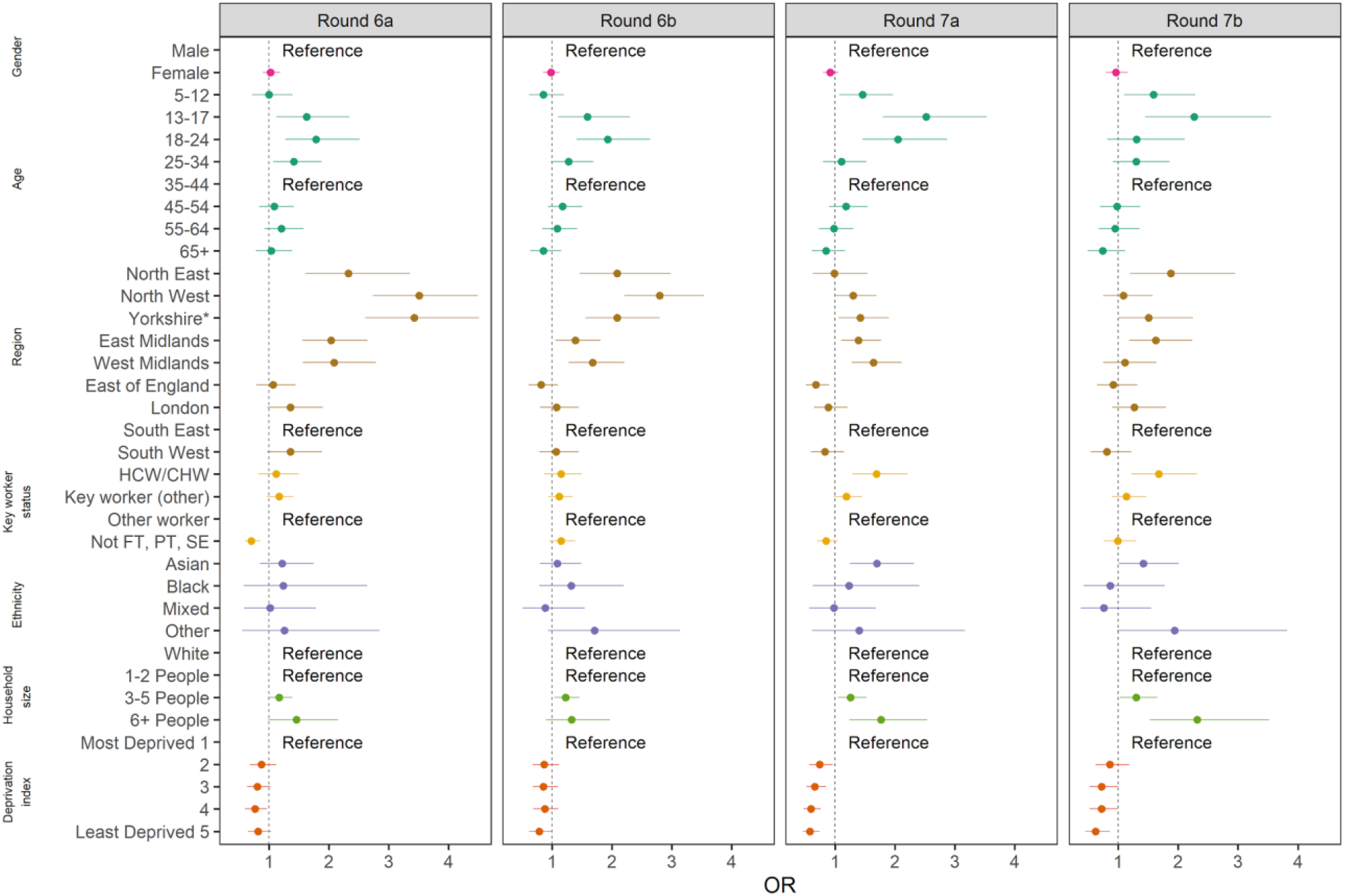
Estimated odds ratios and 95% confidence intervals for multivariable logistic regression model of swab-positivity for rounds 6a, 6b, 7a and 7b. Models were adjusted for gender, age group, region, key worker status, ethnicity, household size, and deprivation index. The deprivation index is based on the Index of Multiple Deprivation (2019) at lower super output area. Here we group scores into quintiles, where 1 = most deprived and 5 = least deprived. HCW/CHW = healthcare or care home workers; Not FT, PT, SE = Not full-time, part-time, or self-employed. *Yorkshire and The Humber.

## Discussion

We report a 28% reduction overall in prevalence between our previous round of data collection from mid-October to beginning of November 2020, and the most recent round that ended 3rd December. National prevalence was ∼1%, similar to rates seen in mid-October 2020. This indicates that -- nationally -- the second lockdown appears to have been effective at reducing rates of infection, although less so than in the first lockdown [9]. However, based on regional estimates of R numbers for round 7, there was geographical heterogeneity, with a decline in West Midlands and growth in London. Furthermore, the previous focus of infection which was in the 18 to 24 year-olds had shifted to school-aged children, especially those in 13-17 year-olds where prevalence was ∼1 in 50.

The recent apparent increase in prevalence in London was not seen uniformly, but was concentrated in the east and south-east of the city with some areas of higher prevalence also seen to the west. In addition, higher rates were found in adjoining areas in East and South East of England (in Essex and Kent respectively). Because of the focus on younger, school-aged children, a programme of rapid testing using lateral flow antigen assays was recently announced in schools in the most affected areas of London, Essex and Kent [14].

Reasons for the differential regional effects of lockdown are not clear but may include the fact that in the northern regions there has been pressure on hospital services due to COVID-19 and large parts of these regions have been in the strictest containment measures through a national tiering system since before lockdown; some of the recent improvements may therefore reflect the combination of these prior measures and lockdown. In London and the south, the rapid rise in prevalence in September to October 2020 previously reported [6,10] has not yet resulted in the very high prevalence rates that were observed in the north, suggesting that, in the absence of lockdown, rates may have gone much higher. However, there is no room for complacency. If rates in London and elsewhere continue to increase, there will be additional strain on the health service in the run up to Christmas.

Our data suggest that on average ∼700,000 people were infected with the virus on any one day of round 7, assuming that the swab procedure in our study detects around 75% of infections [15]. As in previous rounds of REACT-1, we found higher prevalence rates of swab-positivity among people of Asian and Other ethnicity (mainly Arab people) compared with white people and in those living in the most deprived areas. We also continue to report higher prevalence among hospital and care home workers compared with other workers, suggesting that clinical and care settings remain a risk for transmission. However, odds were much lower than seen at the end of the first wave, when hospitals and care homes were major drivers of the epidemic [9].

Our study has limitations. While we include random samples of the population, participation will vary by socio-demographic factors which may introduce bias into the prevalence estimates. Since it is possible that people with riskier behaviours with respect to transmission of the virus may be less likely to take part, we may be under-estimating prevalence despite re-weighting our sample. Nonetheless we have no reason to believe that any such biases have been operating differently over the period of the REACT-1 surveys, so time trends and estimates of R are likely to be less affected than absolute measures of prevalence. We have evidence to suggest that prevalence rates may change rapidly in response to behavioural or other changes, but with associated uncertainty [12]. Although we use statistical approaches to smooth rates over time (splines), we also display the calculated daily rates and confidence intervals, allowing the variability in the underlying data to be assessed. Rates at a regional level are less certain than at national level. Nonetheless, regional splines may provide early warning of increases in prevalence. Taken together with other available data, including data from the national testing programme and hospital admissions [3], our regionally stratified data give a more complete picture of how the epidemic is evolving over time.

In summary, the lockdown in England during the second wave of the COVID-19 epidemic has been accompanied by a reduction in prevalence of SARS-CoV-2 nationally. This has helped to offset the large rises in prevalence observed during October 2020. However, the fall in prevalence during lockdown was not seen uniformly across the country; in particular, we found evidence for a recent rise in London and a flattening off elsewhere. Continued vigilance is required to reduce rates of infection until effective immunity at the population level can be achieved through the vaccination programme.

## Data Availability

Link to daya within manuscript

https://docs.google.com/spreadsheets/d/1NxrqmW1Te2MDV_hrazlr2usPntoeS6B5BltXWTlsvH4/edit?usp=sharing

## Supporting information

Supporting data are available [here](https://docs.google.com/spreadsheets/d/1NxrqmW1Te2MDV_hrazlr2usPntoeS6B5BltXWTlsvH4/edit#gid=1192958503).

## Acknowledgements

SR, CAD acknowledge support: MRC Centre for Global Infectious Disease Analysis, National Institute for Health Research (NIHR) Health Protection Research Unit (HPRU), Wellcome Trust (200861/Z/16/Z, 200187/Z/15/Z), and Centres for Disease Control and Prevention (US, U01CK0005-01-02). GC is supported by an NIHR Professorship. PE is Director of the MRC Centre for Environment and Health (MR/L01341X/1, MR/S019669/1). PE acknowledges support from Health Data Research UK (HDR UK); the NIHR Imperial Biomedical Research Centre; NIHR HPRUs in Chemical and Radiation Threats and Hazards, and Environmental Exposures and Health; the British Heart Foundation Centre for Research Excellence at Imperial College London (RE/18/4/34215); and the UK Dementia Research Institute at Imperial (MC_PC_17114). We thank The Huo Family Foundation for their support of our work on COVID-19.

We thank key collaborators on this work – Ipsos MORI: Kelly Beaver, Sam Clemens, Gary Welch, Nicholas Gilby, and Kelly Ward; Institute of Global Health Innovation at Imperial College: Gianluca Fontana, Dr Hutan Ashrafian, Sutha Satkunarajah and Lenny Naar; North West London Pathology and Public Health England for help in calibration of the laboratory analyses; NHS Digital for access to the NHS register; and the Department of Health and Social Care for logistic support. SR acknowledges helpful discussion with attendees of meetings of the UK Government Office for Science (GO-Science) Scientific Pandemic Influenza – Modelling (SPI-M) committee.

